# A grounded theory study of beliefs underlying use of ancestral spirits for healing among Baganda traditional spiritual healers in Central Uganda

**DOI:** 10.1101/2024.04.03.24304941

**Authors:** Yahaya H. K. Sekagya, Charles Muchunguzi, Payyappallimana Unnikrishnan, Edgar M. Mulogo

## Abstract

The increasing global sensitivity to spirituality in medicine and sociology has elevated the relevance of beliefs in ancestral spirits as an integral element in Africa’s multi-ethnic society and biocultural diversity for health management. However, ancestral spirituality in healthcare in the local context, remains sparse and elusive since most literature relates spirituality with religion and religious practices. The study sought to explore the beliefs in ancestral spirits utilized for health management among practicing Baganda traditional spiritual healers (*Balubaale*). Qualitative data were gathered through semi-structured in-depth interviews and partial integration observation methods from twelve (10M, 2F) purposively selected and recruited *Balubaale* from Central Uganda between 15^th^ July 2019 and 29^th^ April 2020, and we prospectively interacted with them for 24 months. Transcribed data was coded and thematically analyzed using ATLAS ti. 22 computer software based on grounded theory approach. Data revealed themes related to beliefs in spiritual powers of categories of ancestral spirits, sacred places, living plants and animals, non-living things such as fire, water, the Sun, and non-materials such as symbols and colors. Spiritual powers can be potentiated through rituals, sacrifice and communal activities all of which are utilized during health managements. In conclusion, *Balubaale* believe that illness and health management are influenced by ancestral spirits and spiritual powers contained in nature and creation therein. We recommend more exploratory studies among spiritualists of other tribes to contrast the findings.

## Introduction

Every community in a pluralistic society seeks to build her individual health culture based on its traditions (1,2) grounded in sustained beliefs of the people and place (3). The relevance of beliefs in ancestral spirits (4,5) has been raised by increasing global sensitivity to spirituality in medicine and sociology (6–8). Beliefs in health management using ancestral spirits is regaining its grounds as an integral element of Africa’s multi-ethnic society and biocultural diversity (9,10), and the human experience of connectedness with sacred nature (11).

African local ancestral spirituality in healthcare remains sparse and elusive (12,13), since most literature relates African health spirituality with religion and religious practices (14,15). Africans beliefs patterns are rooted in cultural knowledge with personal dimensions to maintain, preserve, restore and heal their bodies and reduce suffering (16). Beliefs provide emotional reassurance favoring hormonal, autonomic and immunologic responses to health problems conceptualized in line with cultural explanations incorporating traditional beliefs (17), metaphysical worldview (18,19), and witchcraft (17,20). Various recommendations have been made to understand the importance of ancestral spirits, ritual healing (8,19) and spirit mediums in health management (21,22).

In Uganda, literature regarding beliefs and role of ancestral spirits in health management is limited (23–25). Therefore, we sought to explore the beliefs and role of ancestral spirits in health management among Baganda traditional healthcare spiritualists (*Balubaale*) from Central Uganda. The research question addressed to participating spiritualists was “What beliefs guide you in your health care practice?”. This paper does not intend to discuss traditional healing system nor evaluate its effectiveness.

## Method

### Study design and Setting

We used grounded Theory (GT) to inductively create and build a substantive theory around the beliefs Baganda traditional healthcare spiritualists (*Balubaale*) base on to engage ancestral spirits in health management (26). Participants created meaning through communication with us, the researchers. We explored, reflected, and conveyed the participants’ meanings in the emergent theory (26). We reviewed some literature to conceptualize and set the scope of study, but avoided extensive literature review before the study to ensure theoretical sensitivity (our ability to detect meaning in the data) (26). Data was collected from 11 districts of Central Uganda, originally inhabited by Baganda ethnic group and Luganda is the main local dialect spoken by the people.

### Participants and sampling

We recruited 12 (10Male; 2Female) experienced Baganda traditional healthcare spiritualists (*Balubaale*) between 15^th^ July 2019 and 29^th^ April 2020 and prospectively interacted with them for two years. We selected *Balubaale* who engage ancestral spirits during health management, with more than 10 years of experience and had trainees at their shrines. Participants were identified by leaders of their associations, districts and clans, and were purposively selected to include Baganda spirit mediums who worked and resided in Central Uganda. Adjustments were done to include *Balubaale* mediums for ancestral spirit “Muwanga” because it was established during field study that spirit mediums for ancestral spirit *Muwanga* were ethical and genuine in the practice. Only 12 participants fulfilled the inclusion criteria out of the original list of 149 names (See figure 1). The study excluded spiritualists who confessed to be of other tribes and those residing outside the study area (27).

**Figure 1.**
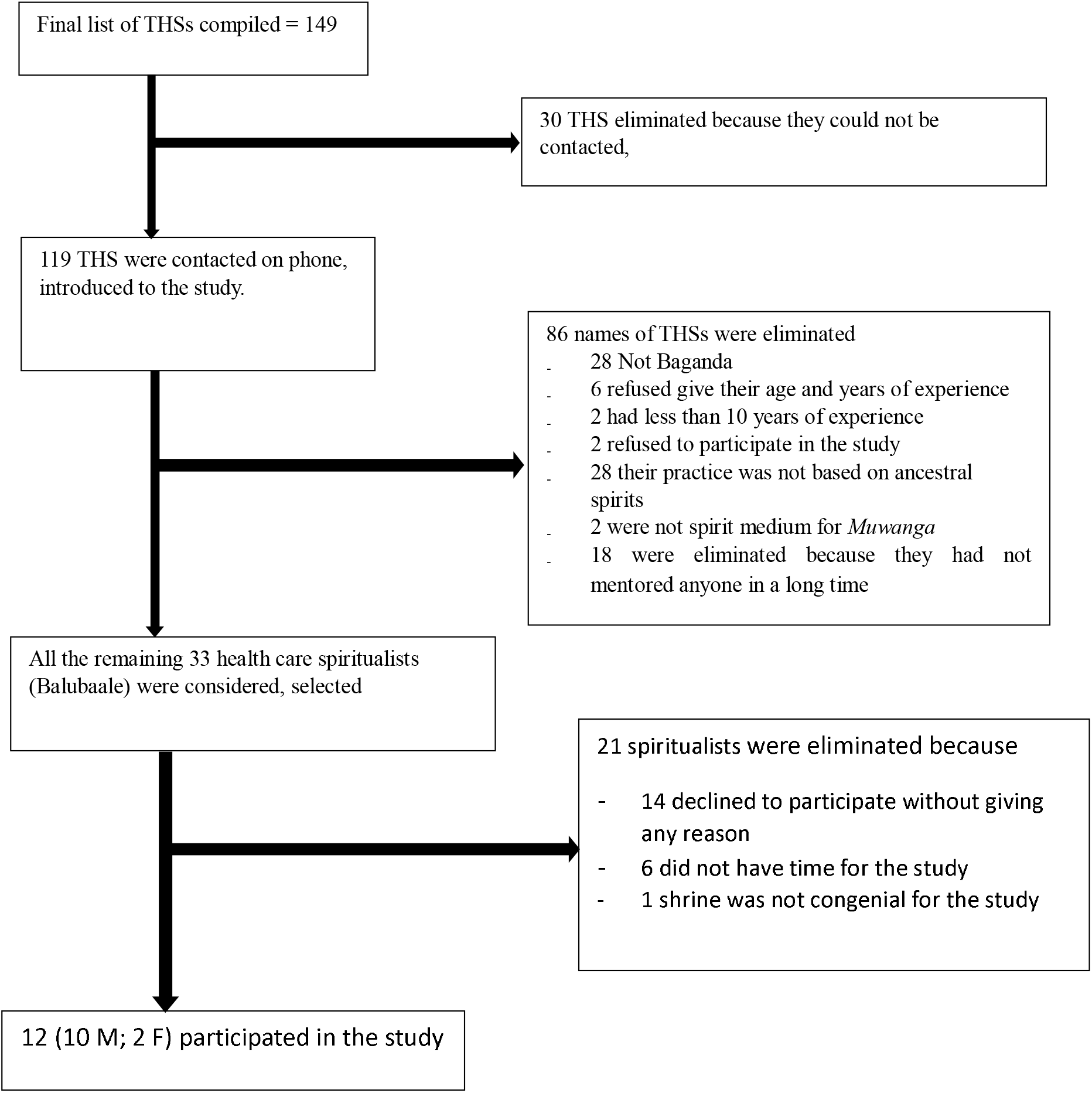
Study profile.

### Data collection and management

We conducted in-depth face-to-face unstructured interviews using open-ended questions and partial integration observation methods to capture and understand the beliefs underlying the health management practice (28) of *Balubaale*. The guiding question was “What beliefs guide you in your health care service provision?” The probing questions related to cultural, spiritual, supernatural and metaphysical forces, the ancestral spirits, and spiritual powers of various materials and non-materials mentioned by the participants, including plants, animals, stars, symbols, regalia and rituals among others. More revealing insights were got in discussions that continued after formal interviews where we flexibly provoked subjective beliefs and worldview of their social reality and contextual meaning (29,30).

The first author revealed his orientation and personal involvement in the research (31,32) and the research team was involved in daily healthcare activities which lessened interference with their usual practices. The research team was trained in note-taking, personal self-reflective journal (33), guiding questions were reviewed by expert in qualitative research, translated in local dialect Luganda and pilot tested with two *Balubaale* in Kyagwe and Busiro counties. Conversations were audio recorded and conducted in Luganda local language of research participants and research team. Recorded audios were transcribed in English while maintaining depth of context in Luganda. All transcripts were reviewed by research team, independently coded by researchers, results compared, merged, agreed upon and used by the research team throughout data coding. Collected and transcribed data was duplicated and progressively analyzed using ATLAS.ti 8, upgraded to ATLAS ti. 22 computer software. Initial analysis of responses was done after first three sets of unstructured interviews and subsequent interviews were compared with previous. We frequently returned to previous participants to capture more data and clarify over some already captured data. Distinct inconsistence and disagreements were redone and cleared by research team.

## Data Analysis

We used the following steps during data analysis shown in figure 2 below

**Figure 2.**
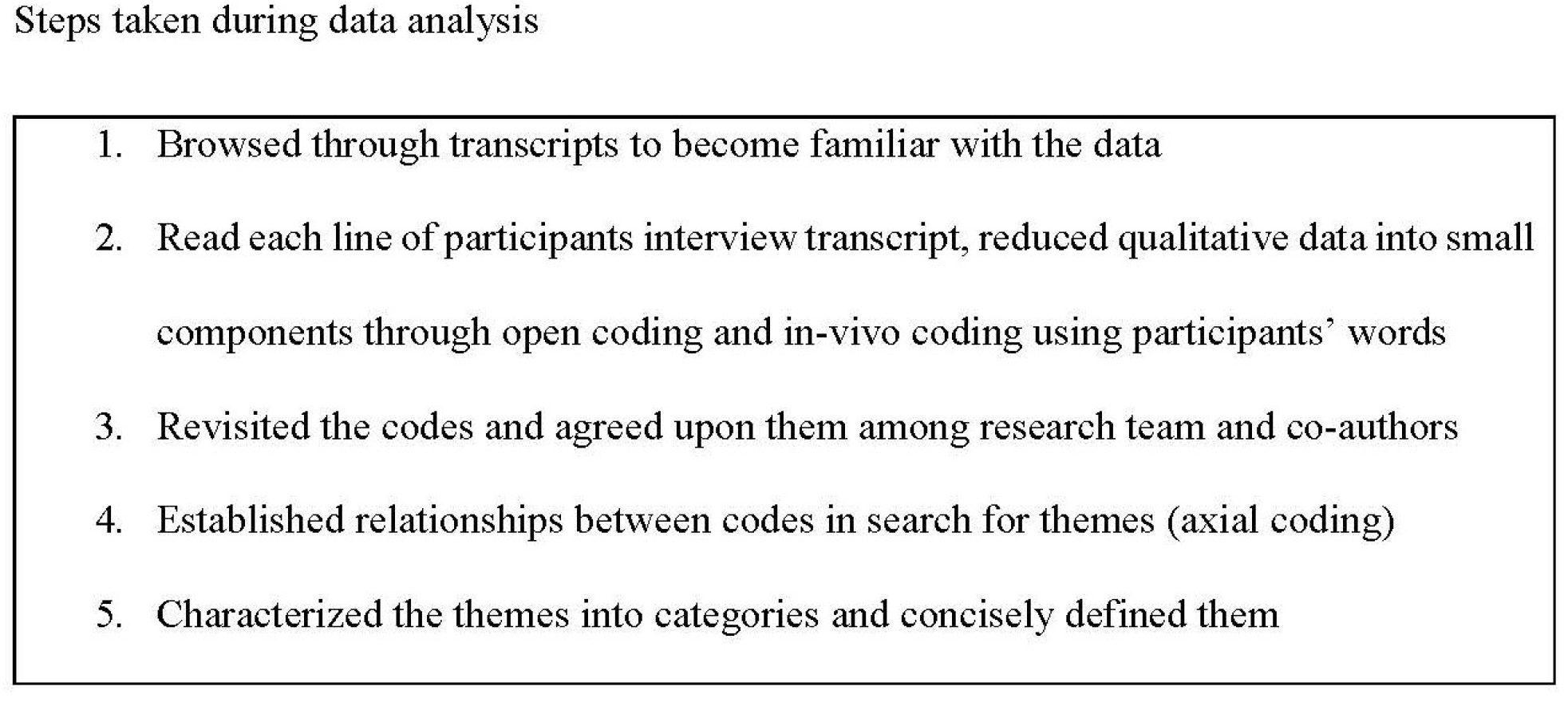
The steps taken during data analysis.

### The 5 steps taken during data analysis

#### 1. Browsing through transcripts to become familiar with the data

Familiarizing with data started during data collection and continued through data transcription. During transcription, time codes were inserted in transcripts to enable revisiting vital parts of conversations for clarity and in case of need for additional interviews. Any disagreements in transcriptions were discussed by research team, the original audio-recording revisited, and at times, responsible participants were consulted for clarity.

#### 2. Line-by-line reading of each of the participants interview transcript

Each transcript was read many times by all individuals of research team. Through this in-depth reading, we wrote the ideas of patterns. After obtaining overall understanding of the data, we created an ATLAS.ti project and added documents as they were developed and each document was automatically assigned a number. (Figure: *List of documents with meta information*). Qualitative data was reduced into smallest components in form of codes (open coding). Some codes were given names using participants’ words (in-vivo coding) (34). Initial coding process labelled the topics mentioned by research participants and observational notes to describe the data.

#### 3. Revisited the codes and agreed upon them as research team and co-authors

Coded segments were validated and agreed upon by research team to find meaningful headers to code and organize the codes with aim of building a final code structure. Code frequency in the Code Manager, was instrumental in renaming, merging some codes with low frequency or splitting up of group codes with very high frequency. The codes in the first code structure were defined to clarify their exact meaning and application.

#### 4. Established relationships between codes in search for themes (axial coding)

After refining the codes, similar codes and concepts were compared, grouped into meaningful pattern, linked and organized by relationships to form themes (35,36). Themes were reviewed and discussed with co-authors. Memos were written for each theme and linked to data segments that illustrate main aspect of the theme.

#### 5. Characterized the themes into categories and concisely defined them

The agreed upon themes were linked to make meaning, named, and their networks diagrammatically visualized using ATLAS.ti software. We then described core essence of each individual theme in a concise and informative manner (37). We related themes to each other in a process that involved moving between themes, collapsing some while expanding others.

Data was analyzed using inductive approach. Epistemologically, the research was rooted in interpretive approach, where reality is considered subjective, multiple and socially constructed. The main results of analysis were to give an interpretative understanding of *Balubaale’* beliefs through experience of their reality as shaped by their historical and social-cultural perspectives (36). By the time of the 10^th^ participant, much data was repetitive and had reached data saturation. The whole research process was not linear, as we moved back and forth through the process (38). All efforts were made during data interpretation to remain close to the context of the relationships and scenarios in which information was gathered.

### Development of substantive theory grounded in participants data

We use the following phases during data synthesis using ATLAS.ti 22

**Figure 3.**
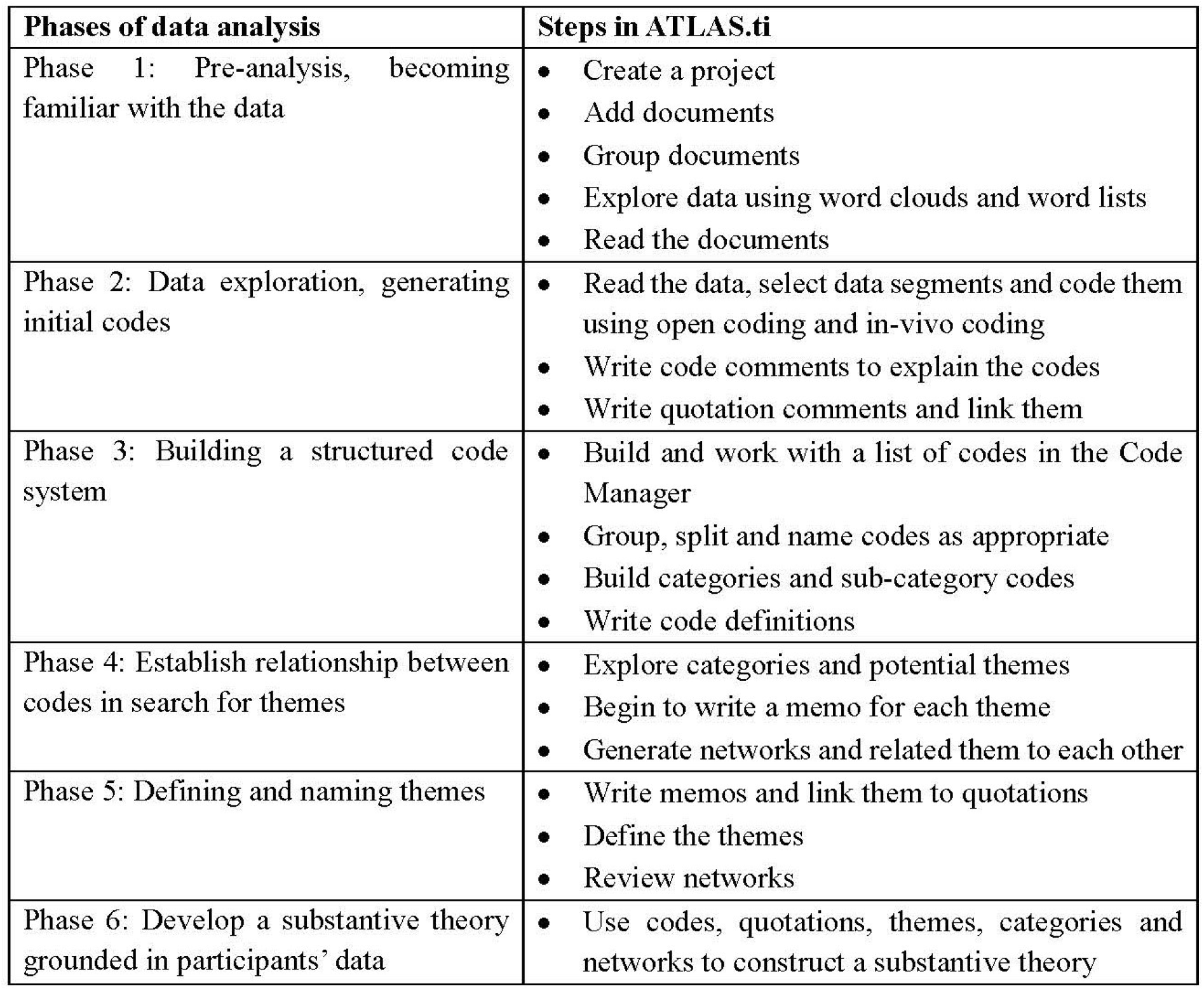
Table showing phases within ATLAS.ti 23 data synthesis.

After development and discussion of the emergent theory, we returned to the original research *Balubaale* participants at their shrines, to validate and confirm the theory.

### Ethics consideration

Ethical approval of this study was sought and obtained from the Research and Ethics Committee at Mbarara University of Science and Technology (No. MUREC 1/7) and the Uganda National Council of Science and Technology (SS 4947). The study was also cleared by Traditional Healers’ Associations (NACO/0485/2019), and Buganda Kingdom. Written prior informed consent was also obtained from the individual subjects.

## Findings

### Socio-demographic characteristics of respondents

Study participants were aged between 27-77 years (Mean age 54), from 8 counties, 11 Districts, and 9 clans. 8 belonged to traditional religion. Two of them never attended school, 6 never completed primary level and 4 completed secondary level education. All respondents belonged to healers’ associations, had more than 10 years of experience and were subsistence farmers. Two doubled as spiritualists using ancestral spirits (*Balubaale*). and spiritualists using nature forces (*Baluutansoz*i). Three participants started spiritual healing when they were less than 18 years old. The socio-demographics of the study participants are represented in the figure 4 below.

**Figure 4.**
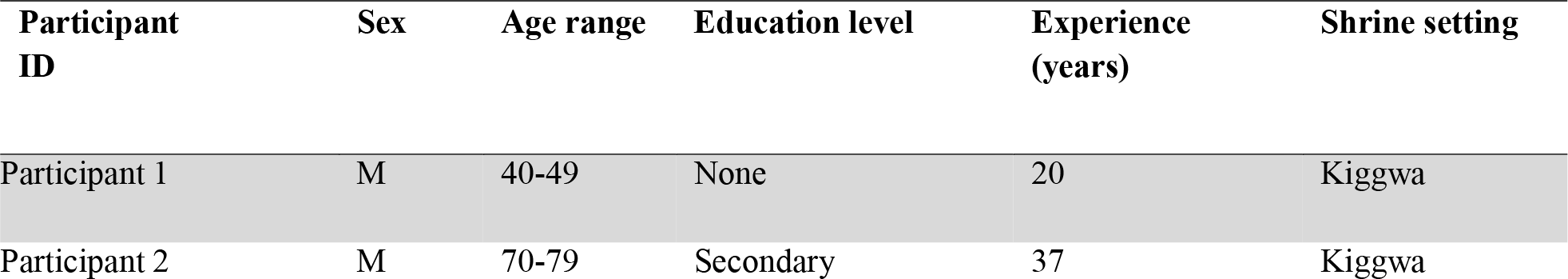
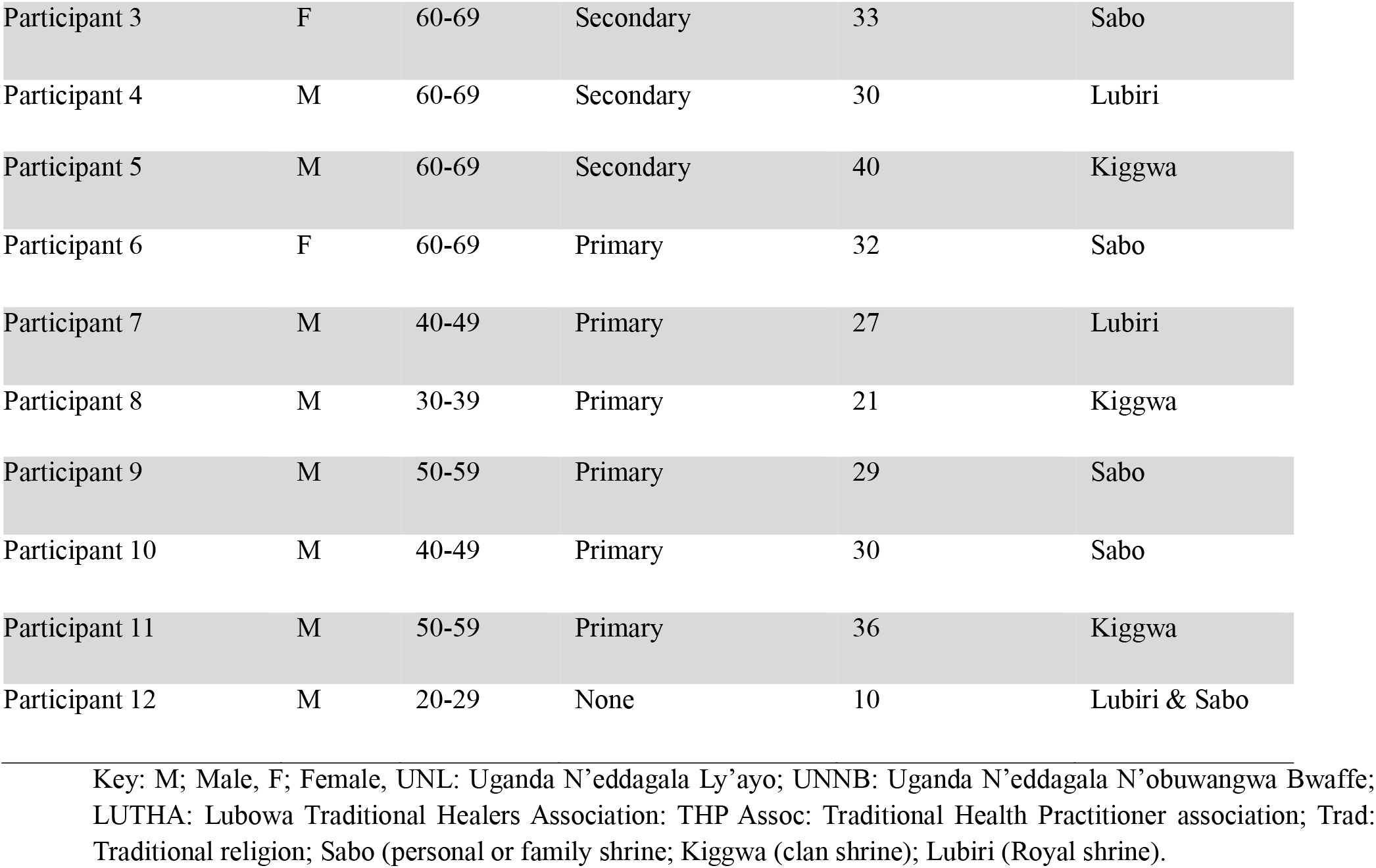
Table: Visualization of Socio-demographics of study Participants.

Central Uganda is Buganda, and is comprised of 18 counties, 24 districts and dominated by Baganda ethnic tribe with 56 clans (39). This implies that the study sample is not representative of clans of Baganda in Central Uganda. Traditional spiritual healing is enshrined in socially and culturally constructed community gender specific taboos, and is historically dominated by men. However, women are reported to be comparably greater in reproductive and domestic issues (40). In this study, being a medium for male ancestral spirit *Muwanga*, was part of the inclusion criteria of respondents. These gender specific restrictions may partly explain the fewer female participants in the ratio of 5:1 of men to women.

### Traditional beliefs in ancestral spirits

Baganda traditional spiritual healers (*Balubaale*) believe in ancestral spirits and spiritual powers contained in sacred places, spiritual materials, and rituals. Spiritual materials include living and non-living things, and the spirits were expressed in two broad categories, natural and ancestral spirits. They also believe that ancestral spirits have inheritable experiential knowledge and experience, are responsible for causing some illnesses and are used in health management. This article has focused on participants beliefs in three categories of ancestral spirits that work through spirit mediums (*Lubaale asamirwa)* namely; spirits of the dead persons (*Mizimu*), mystical ancestral spirits (*Misambwa emizaale*) and the worker/assistant spirits (*Mayembe*), as reflected in the figure 5 below.

**Figure 5.**
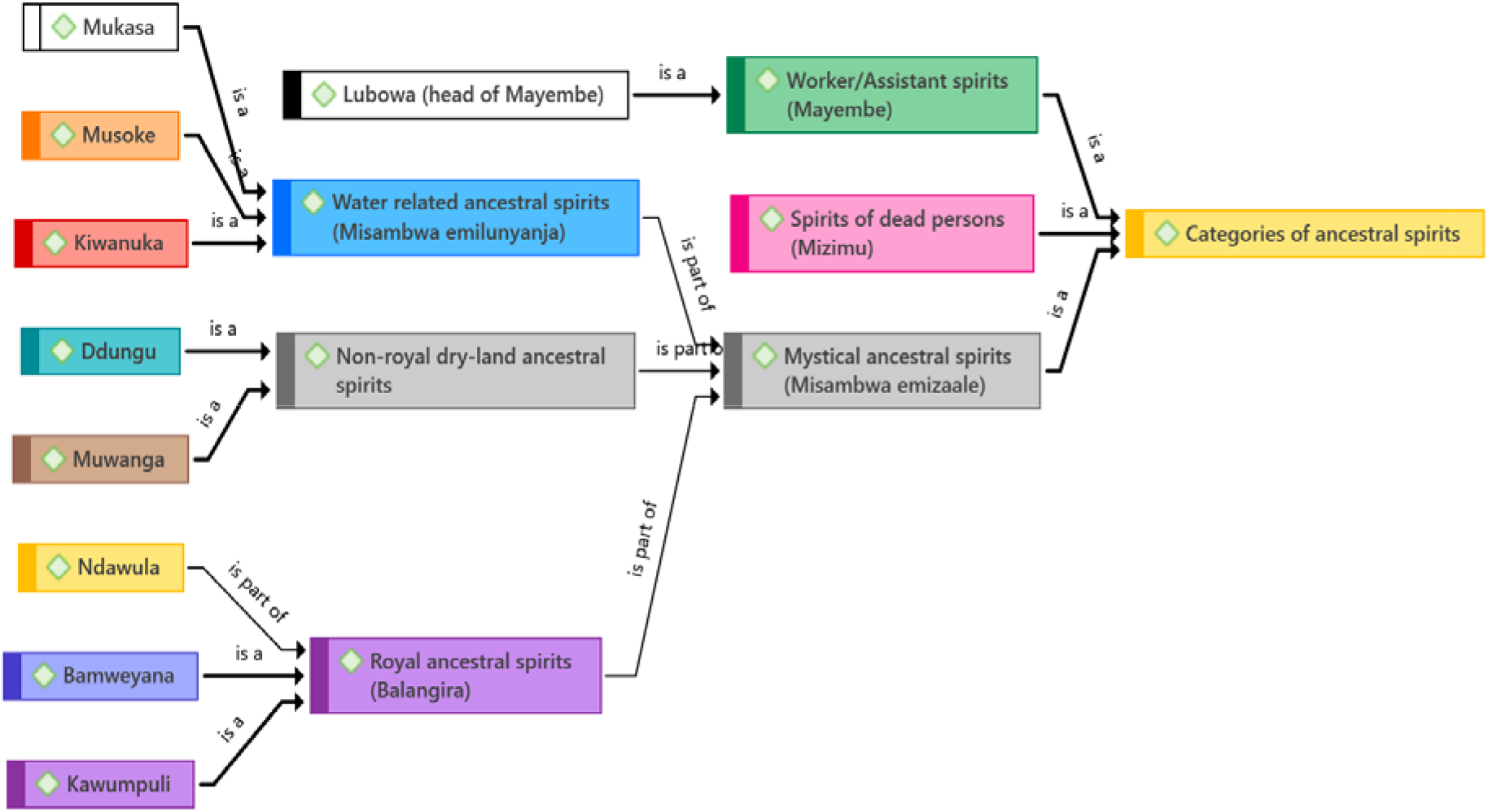
Diagram showing the categories of ancestral spirits considered in this study.

### Characteristics common to *Misambwa, Mizimu* and *Mayembe*

The mentioned characteristics below apply to all these categories of ancestral spirits. They are respected, have earthly activities and often select their human mediums. They frequently communicate through dreams and interact with human family and clan members for whom they offer protection and maintain health. Each category of ancestral spirit has particular regalia embedded with spiritual powers that are also utilized during health management. However, when repeatedly offended, they care be responsible for the cause of illness and subsequent diseases.

Each category is able to engage in health management activities such as divination, health assessment, harvesting of medicines, preparation, prescriptions, dispensing and performs rituals. They have sets of diagnostic tools comprised of various natural materials such as cowry shells, rocks, plant materials, animal and birds’ parts, etc. Each category of ancestral spirit has particular characteristics below.

### Characteristics of spirits of the dead persons (*Mizimu*)

Participants described the spirit of dead person as *Muzimu* (*Mizimu*-plural) to have authority over all other ancestral spirits. One participant expressed that “*Muzimu (name withdrawn) is an ancestral spirit of my great-great grandfather for which I am a spirit medium”* (Participant 1). After a very long time, the *Mizimu* may transform into mystical ancestral spirits (*Misambwa*).

### Characteristics of mystical ancestral spirits (*Misambwa emizaale*)

Mystical ancestral spirits (*Misambwa emizaale)* are found in all Baganda clans and are believed to be among the eldest and most superior ancestral spirits whose functions are controlled by *Mizimu.* They work directly with strong spiritual natural forces. *Misambwa emizaale* are sub-grouped into three, namely; water related ancestral spirits (*Misambwa emilunyanja),* royal ancestral spirits (*Misambwa emilangira)* and non-royal dry-land ancestral spirits (*Misambwa jo kulukalu)* such as Dungu and Muwanga*. Misambwa* are associated with spiritual powers of sacred places, plants, animals, waters and colors. The role of *Misambwa* in health management relate to removing or eliminating the root causes of illness and manifested diseases. These spirits are sometimes jointly referred to as *Lubaale we Nyanja,* and are believed to have originated from Ssese Island across Lake Victoria locally called *Nyanja Nalubaale*.

### Water related ancestral spirits (*Misambwa emilunyanja*)

The three most mentioned water-related mystical ancestral spirits were Mukasa, Kiwanuka and Musoke. There are closely related and rituals are usually performed at about 5.00 am. Mukasa, Kiwanuka and Musoke are biological grandchildren to Kabaka Mukasa Sebyoto.

### Ancestral spirit Mukasa

Mukasa essentially refers to two ancestral spirits, The eldest Kabaka Mukasa Sebyooto Lukankana, and Mukasa the grandson. Both Kabaka Mukasa and the grandson Mukasa have similar characteristics with the following variations. Kabaka Mukasa Sebyooto Lukankana expresses himself as a very old man with shaky voice, trembling body and hands. He is highly experienced and works as a health consultant. He mostly delegates his works to other spirits. While Mukasa, the grandson, presents itself as an adult man with strong voice and body.

Both express their presence by symbol of sailing in a canoe boat using an oar or paddle. They great using “*Ssese Davu – gayira Ssese gayira”*. Both do not use alcohol but instead use water, fresh juice especially from ripe banana, and honey. They use a fire-place called *ekyoto kya Mukasa*, and their offering does not involve pouring of blood. They both work mainly during day time. They are specifically associated with good health, good luck, blessings for individuals, families and communities. Both help to address infertility in humans. For example, two spiritualists said; *“… at my shrine, Kabaka Mukasa is head of all ancestral spirits responsible for health assessment and diagnosis, offers good luck and heads the healing rituals.”* (Participant 3). *“Mukasa is the ancestral spirit that confirms issues regarding ancestral spirits and their activities.”* (Participant 12). Ancestral spirits Mukasa are associated with white and silver colors, that is white animals and birds, and white and silver artefacts and regalia.

### Ancestral spirit Kiwanuka

Ancestral spirit Kiwanuka was described as a fast-acting (swift), practical spirit with few words, but talks with a strong, loud, forceful voice. Kiwanuka is characterized by holding, eating or stepping on live fire while his medium is dancing to rhythm of loud songs and drums. Kiwanuka is symbolized with red and brown colors for its artefacts, regalia, and animals or birds. Most of its rituals are associated with use of brown, uncastrated male sheep, goat, bull and a brown rooster (*nkoko ya lujuumba omumyufu*). For example, two participants said *“during my initiation, ancestral spirit Kiwanuka demanded for a brown, adult male sheep, and an adult male brown rooster chicken.”* (Participant 1). Furthermore, Kiwanuka is also associated with spiritual energies of mountains, fire, the Sun and the plant ficus branchypoda locally referred to as *Mukokowe or omuserere*). One participant made a comment while pointing at a plant *“… this plant (Ficus branchypoda) helps ancestral spirit Kiwanuka to descend in full spiritual powers. It just planted itself here”)* (Participant 7). Kiwanuka is known for causing illness involving mental disorders such as madness and its management. *“Ancestral spirit Kiwanuka causes madness in some people when not well harmonized”* (Participant 1).

The regalia for ancestral spirit Kiwanuka include spiritual head gear (*Ekisingo*), brown beads, spear and harmer (*Nyondo ya Kiwanuka*) made of copper, and a seating mat (*ekiwu*) made from sheep skin sacrificed for it during harmonization rituals. Others items include brown bracelets, bark cloth made from ficus branchypoda plant, specified red clay pot, a brown “spear” decorated of brown rooster feathers (*effumu lya Kiwanuka*) among others.

### Ancestral spirit Musoke

Musoke is associated with golden-yellow color, water, honey, and fresh banana juice. His regalia is mainly in golden-yellow colors and its clay pot is golden-yellow. His harmonization utilizes a female goat with a patch across its belly (*Embuzi ya luyina*) and a particular type of chicken (*Nkoko ya lusubi*). Participants associated the ancestral spirit Musoke with gynecological, reproductive health, and issues of menstrual cycles for ladies, the spiritual powers of the rain-ball, the Sun, sky and major water bodies. They said that Musoke causes illnesses associated with dehydration, anemia and infertility. Musoke is not directly involved in health management but works through rituals that may involve communal meals (*ebijjulo*) to make necessary corrections that uplift the causes of illness. One participant commented *“Musoke will use symbolic artifacts, rituals and communal meals to uplift witchcraft or curses responsible for illness.”*(Participant 1)

**Figure 5.**
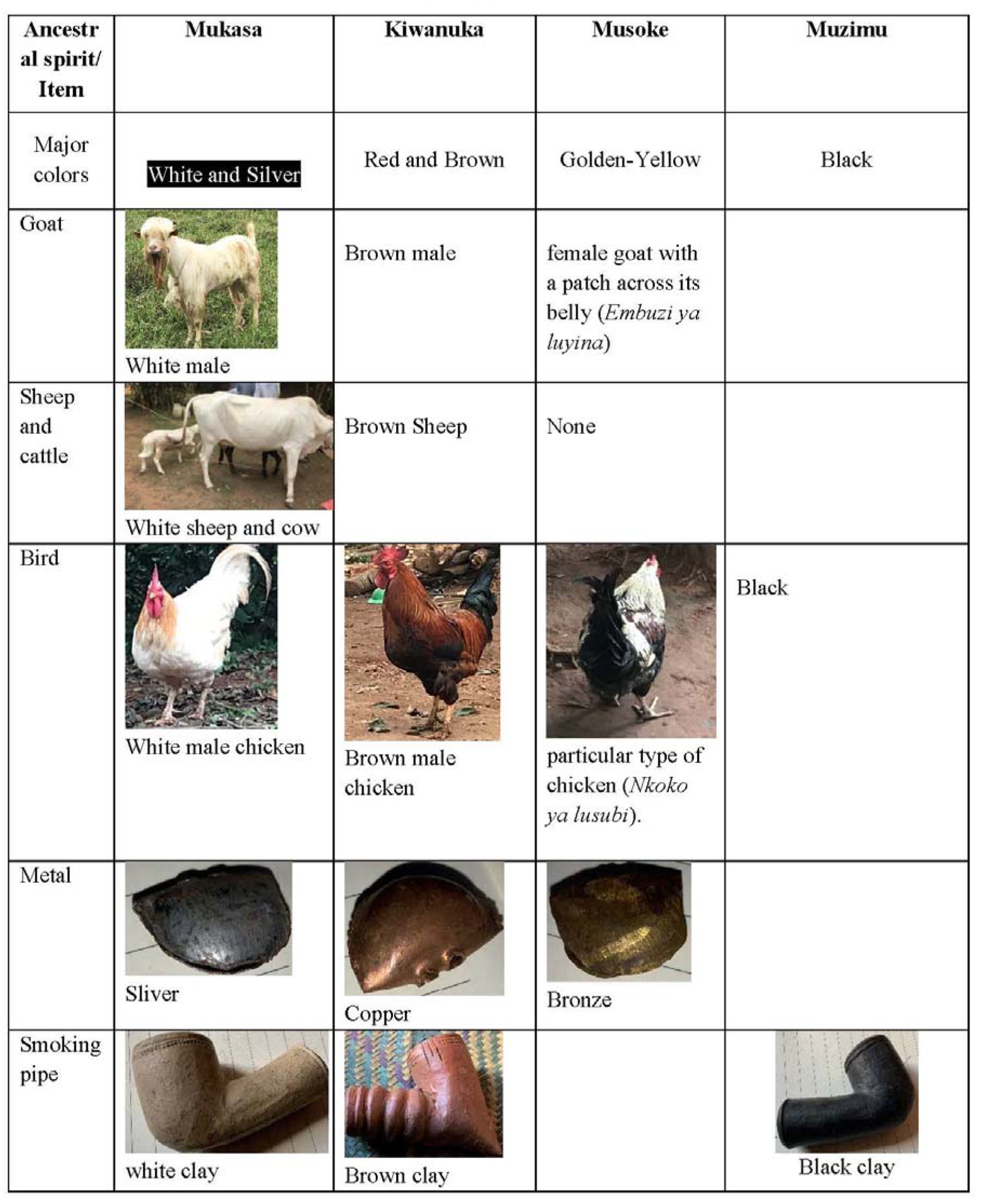
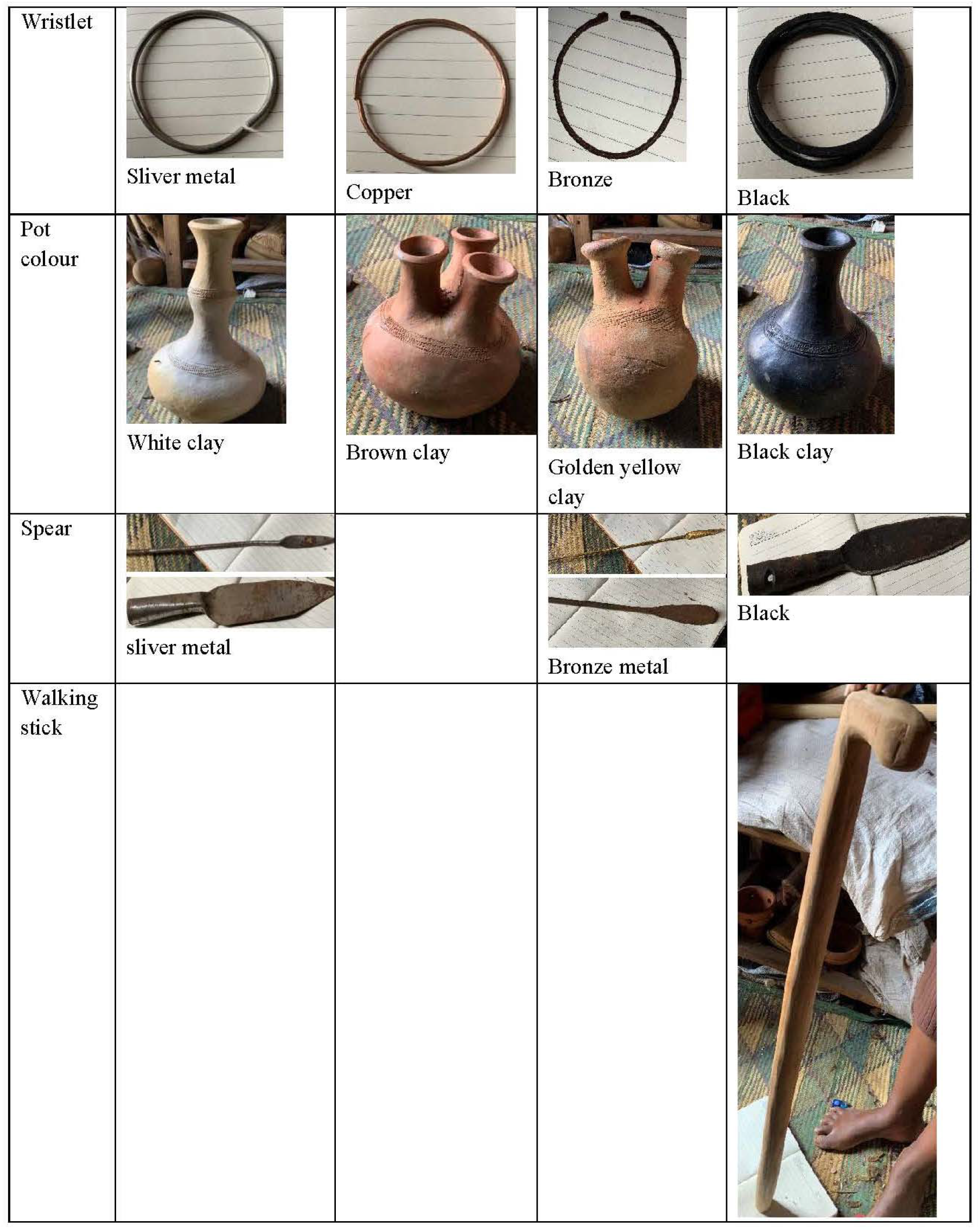
Artefacts, animals and birds associated with *Muzimu* and water related ancestral spirits Mukasa, Kiwanuka and Musoke (*Misambwa emilunyanja*).

### Royal ancestral spirits (*Balangira*)

Royal ancestral spirits *(Balangira)* are the spirits of dead persons of royal origin and are not restricted to any particular family or clan among Baganda. They can possess any person from any clan as spirit medium and are not associated with animal sacrifice during healing. The three most mentioned royal ancestral spirits engaged in health management are Ndawula, Kawumpuli and Bamweyana.

### Ndawula

Participants referred to Ndawula in two spiritual categories, as a royal (*Mulangira*) or as a King (Kabaka). In childhood, Ndawula as a m*ulangira* experienced multiple illnesses, before he later become a King. Ndawula’s shrine is made using dry banana leaves and is offered juice made from fresh ripe bananas, or fresh fruits or honey. His harmonization rituals are done at 3.00 am and require a black or white uncastrated male animal. Ndawula’s artefacts are items with protrusion surfaces including gourds, walking stick, and smoking pipe. His symbols are green and yellow colors

Ndawula is believed to cause individual, family or community problems, illnesses associated with multiple wounds, foul smell and jiggers, warts, rough skin, body swellings, eye pains and cataract all of which may respond to rituals involving materials with physical protrusions. For example, one participant narrated *“when a community destroyed my shrine for Ndawula, the whole community including people, animals and birds were infested with jiggers and lice, not until the community people assembled, apologized, and mobilized themselves to reconstruct the shrine did the jiggers and lice stop in the village”* (Participant 1). Another participant added *“Spirit Ndawula tormented my brother by making his nose rot with foul-smell and non-responsive the western medicine and herbs.”* (Participant 6).

### Kawumpuli

Participants believe that Kawumpuli was born a royal crippler (*Kintuntu),* with special spiritual powers and abilities, by King Kayemba and Princess Nakku. Kawumpuli is referred to as the Prime Minister of all ancestral spirits (*Katikiro w’empewo*), and is referred by many names. His artefacts include a shield (*engabo),* hammer (*enyondo ya Kawumpuli),* and a black spear made of feathers of male adult chicken (*Lujuumba omuddugavu)*. His is offered animals that are reared but not sacrificed. His roles include training and harmonizing other spirits and duties assigned to him as an assistant to Spirit Muganga, whom he also empowers. His specialty include divination, health assessment, diagnosis, treatment, cleansing rituals and training.

### Bamweyana

Participants believe that Bamweyana is a royal promiscuous spirit, often seemingly tainted to be drunk of crude alcohol (waragi), use drugs like marijuana and is associated with a hip of burnt or burning rubbish. His character is expressed in his regalia dressings, and hair style. However, Bamweyana is believed to be very wise, with sober, very good and balanced judgements especially in health management as expressed by two participants. “*Bamweyana is dressed in rags and sisal sack.”* (Participant 11). *“Whereas Bamweyana is drunkard, a womanizer, behaves as a mad person and enjoys fighting, but he is known to be very intelligent, hardworking, with knowledge of medicinal plants and skills of health management.”* (Participant 3). However, Bamweyana is believed to cause madness.

**Figure 6.**
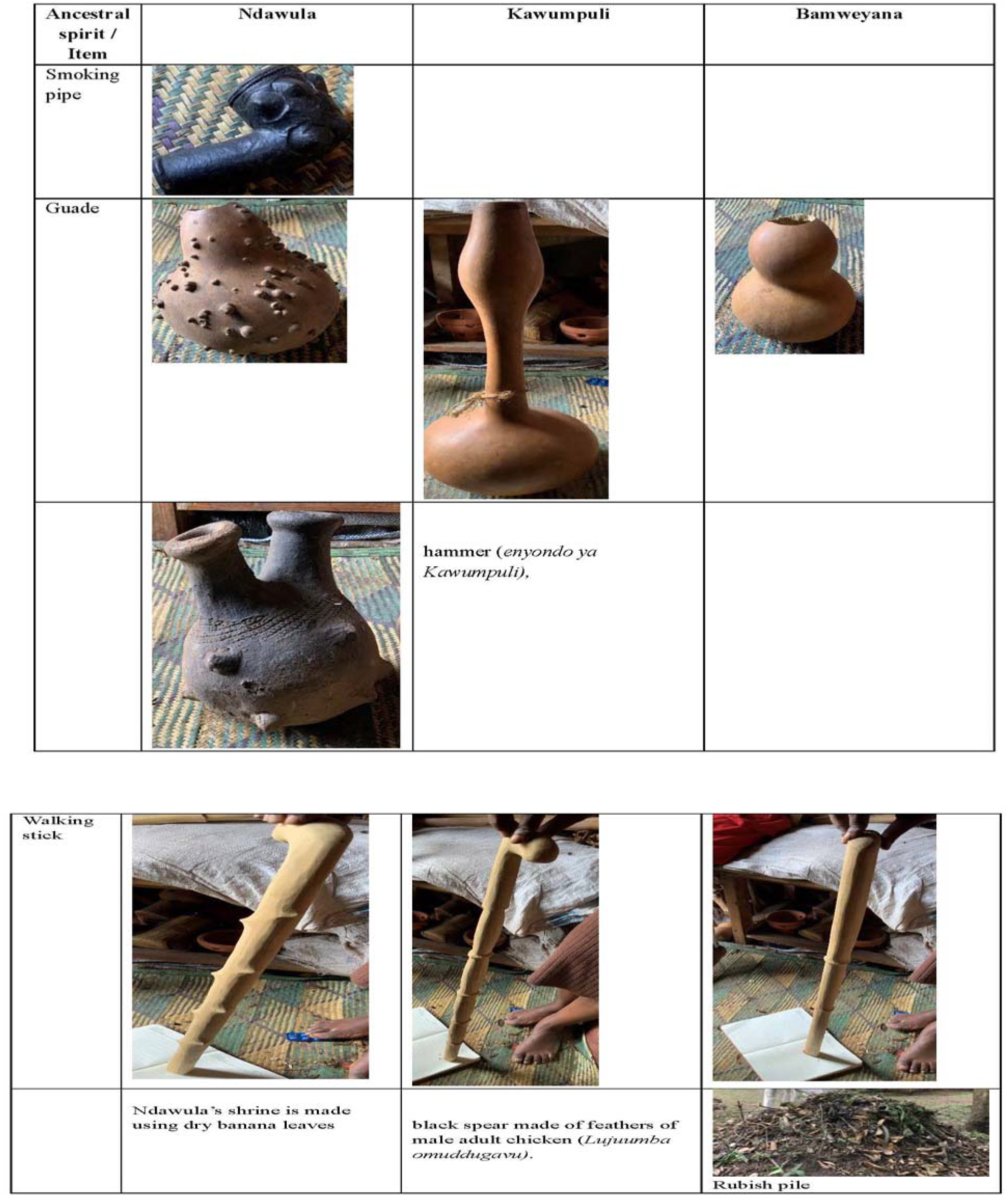
Artefacts associated with royal ancestral spirits (*Balangira*)

### Non-royal dry-land ancestral spirits

The participants mentioned two major non-royal dry land ancestral spirits namely Ddungu and Muwanga.

### Ddungu

Ddungu is considered the chief spirit of the wilderness and is associated with hunting and hunting dogs. His hunting artefacts include a hunting net (*olutuula*), hunting spear (*effumu eliyigga*), ringing bells (*ebide*). Its harmonization rituals are done in the wilderness and require a male, multi-colored goat (*Lumyamyamya)*.

The major role of Ddungu in health management was to collect medicinal materials from the wilderness. When a person was possessed by spirit of Ddungu, they usually go to the wilderness and spend time, picking medicinal plants and or hunting animals which are used as medicinal materials in health management. He has a fireplace (ekyoto kya Ddungu) that is key during health management.

### Muwanga

Muwanga is believed to be the grandparent of all ancestral spirits with unclear origin and without a grave. He just disappeared! Muwanga is multi-phased spirit that manifest either as a young Muwanga from Nseke village (*Muwanga womu Nseke)* or an elderly Muwanga from Kaliggwa village (*Muwanga we Kaliggwa*) in Mawokota county, Mpigi district, Uganda.

Other participants the two Muwanga as one is a grandson of the other. The youthful Muwanga is associated with drinking Alcohol and has a strong voice. The elderly Muwanga, sometimes referred to as Muwanga S*ebyoto Lukankana* is very old, soft spoken with old trailing hardly audible voice. He is hardworking and does not drink alcohol. He is very knowledgeable in the working modalities of nature, very confident in his works and he does not work in a hurry, he takes his time. One participant expressed Muwanga as *“… I am a spirit medium for Muwanga Ssebyoto Lukankana, a very old man indeed, his is not in any hurry, speaks softly but is very confident in his undertakings”.* (Participant 8).

Muwanga’s shrine (*essabo lya Muwanga*) is usually at clan level (*kiggwa*). It has two doors, a fire place (*ekyoto kya Muwanga)* inside it near the entrance. Muwanga’s shrine is used by all ancestral spirits as expressed by one participant *“…Muwanga is a good trainer, and all other ancestral spirits feel at home in Muwanga’s shrine.”* (Participant 6).

Muwanga has a unique, believed to be most accurate diagnostic tool (*omweso gwe ngatto gwa Muwanga),* comprised of nine (9) pieces of various powerful animal hides such as a lion, leopard, hyena, and zebra including a piece of hide of bull sacrificed during its harmonization ritual. It is used by Muwanga as the last resort in any diagnostic inquiry, and Muwanga is considered the best consultant and trainer in all aspects of health management and healing and was a healer of the Kings of Buganda. Muwanga has ability to empower, counsel, train and discipline all other ancestral spirits and empower (*kuwaga*) their respective shrines, regalia and artefacts. Muwanga is believed to be a good psychotherapist, psychoanalyst and an advisor as expressed by one participant *“Muwanga spirit is trusted in his health management style. He is slow but very sure. He explains most of the steps he takes in health management. Muwanga’s ability to calmly explain and patiently answer all questions addressed to him relating to the cause and the approach to the solution is so convincing that it leads the client to build trust and belief in Muwanga”* (Participant 10).

Artefacts for Muwanga include knotted peace of bark cloth (*Kifundikwa kyolubugo*), walking stick, a particular knife (*Omwambe*), cloth robe (*Kanzu*), spear with one or two blades, harmer (*enyondo*), smoking pipe (*emindi)* with one or two heads mostly associated with brown colors.

**Figure 7.**
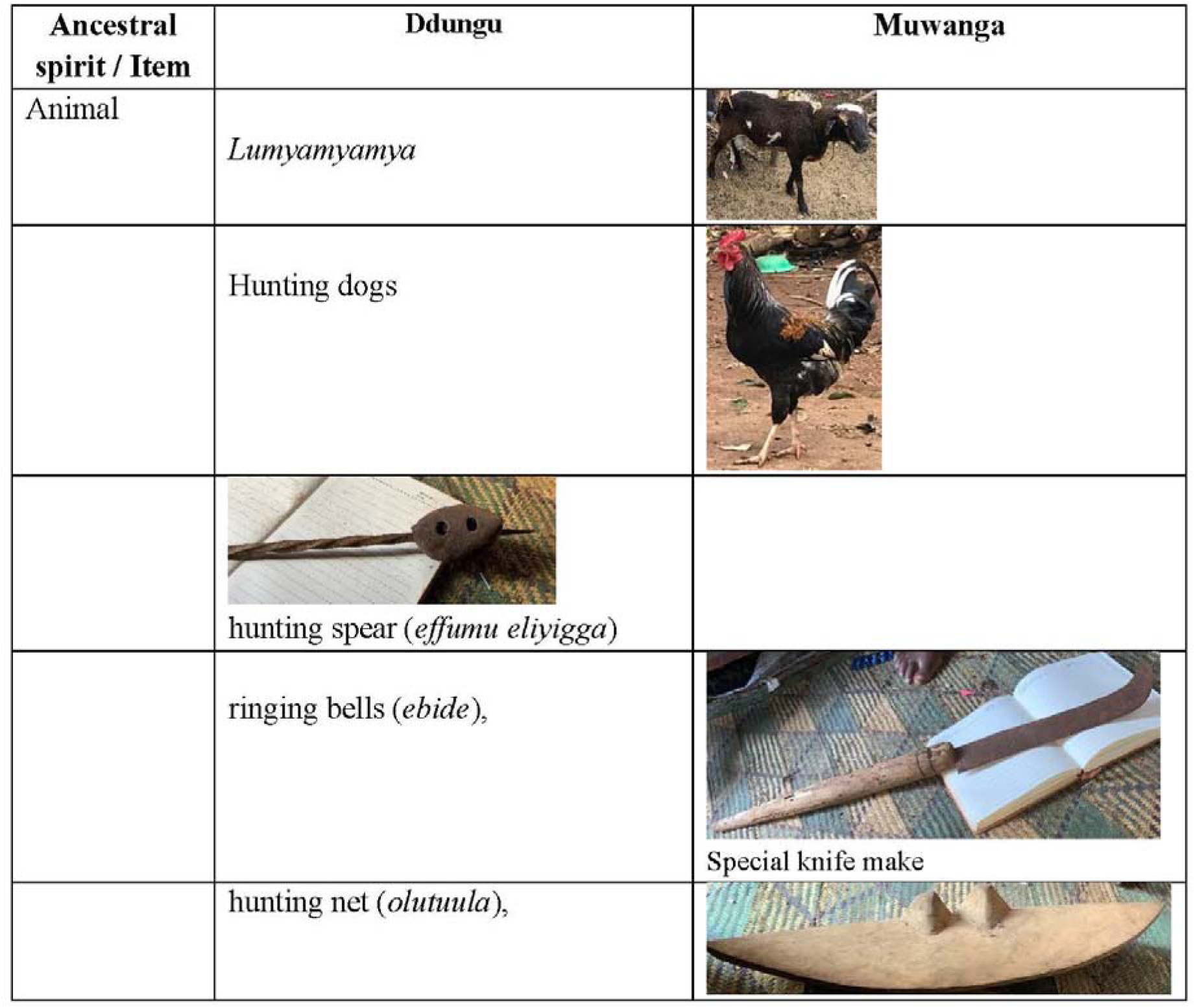
Artefacts and animals of non-royal dry-land ancestral spirits Muwanga and Ddungu.

### Mayembe

*Mayembe (singular. Jjembe*) are categorized into natural, and ancestral or man-made spiritual entities. *Mayembe* trace their origin to Bunyoro Kingdom from where they were brought and adopted as soldier spirits to assist in the war between Bunyoro and Buganda Kingdoms. *Mayembe* that have undergone generations of ritualized harmonization were considered in this study because they were the ones mainly used by *Balubaale* during healing.

Mayembe are protective spirits for other ancestral spirits, people and property. Well harmonized clan *Mayembe* are given strict instructions, guidance, and counselling to perform specific functions during healing, protection of business, family, fight wars, reproduction, upbringing of children and grandchildren. Many Mayembe mentioned included Lubowa, Namuzinda, Kalondoozi, Kasajja and Katabaazi. However, we focused on *Jjembe* Lubowa because participants mentioned it as the head of *Mayembe* ancestral spirits and assigns health management duties to the rest. More details are excluded to shorten the article.

### Other beliefs associated with healing by Baganda spiritual healers

*Balubaale* believe in spiritual healing powers contained in sacred natural and cultural places such as forests, water bodies, mountains, shrines, and fire places (*ebyoto*), living natural materials such as plants, animals, birds, and natural non-living things such as water, rocks, the Sun, Moon and Stars. Such places and items are believed to be dwelling and congregation places for the accessible ancestral spirits, especially the mystical ancestral spirits (*Misambwa*).

### Beliefs in spiritual powers of sacred natural and cultural places

A human spirit medium is required to go to natural sacred places to enhance their spiritual powers, connectivity, recognition, and be able to access and use associated rituals and prayers for healing. Most mentioned sacred places were Tanda, Nyiize, Nalubaale, currently known as Lake Victoria and its Ssese Islands, twin rivers of Mayanja and Ssezibwa, the rocks of Walusi in Bulemezi, and the hill of Misindye in Mawokota. Such places have restricted access and use by initiated spiritual healers and the public. However, the participants sadly noted that most of these places have been adulterated or destroyed and abandoned by the ancestral spiritual entities.

### Beliefs in spiritual powers of shrines and fire places

Appropriately located and constructed shrines and fire-places (*ebyoto*), empowered by rituals performed by spirit Muwanga, were expressed as congregation places for ancestral spirits and their interactions with human beings. These places serve for harmonization, training, empowering, appeasing, and periodically rejuvenating the healing powers of ancestral spirits. Normally the ancestral spirits request for the shrine and give specific requirements for its constructions, including the sacrificial animals and rituals to perform. For example, one participant correlated, *“… the ancestral spirits requested us to construct three shrines, one for Muzimu, the others for Muwanga and Lubowa.”* (Participant 10). The middle pillars are usually of platycalyx (*Musambya)*, Grewia simillis/mollis (*Lukoma)* or Sapium ellipticum (*Muzanganda*) trees. Detailed categories of shrines and fire-places have been not to lengthen this article.

### Beliefs in spiritual powers of living things such as plants, animals and birds

*Balubaale* believe that living things such as plants, animals, and birds have or are associated with spiritual powers responsible for healing. However, when probed further, participants did not find it important to disclose how plants work, but more interested in getting the expected results. For example, one participant explained *“… plants do not work because I can explain how they work, No! Plants work because they have that ability and they give me the expected results. It is not my role to explain how they work. But you may ask the ancestral spirits themselves.”* (Participant 6). The ritual details of harvesting, processing and dispensing for effective healing have been removed on request by the spiritual healers.

### Beliefs in spiritual powers of non-living things such water, Sun and Moon

The participants believed that water is associated with ancestral spirit Mukasa and its healing values are believed to depend on its source such as spring, well, rain, river, lake, or sea/ocean and the associated cleansing and healing rituals. Similarly, the healers believe in the universal celestial powers of the Sun, Moon and Stars which they utilize for divination, healing and guidance. Some healing rituals are done during day while others are done at night. They also believe that the Moon cycle influenced body mechanisms such as menstruation, fertility and reproduction, mental health, mind and intuition. One participant said *“… I use the time of no Moon to pray and fill up my spiritual powers”* (Participant 10). Most ancestral spirits and spiritual powers are associated symbols and symbolism.

### Beliefs in spiritual powers of associated with symbols and symbolism

Participants believe that spiritual energies are represented through symbols such as nature, shape, and color of materials in form of artefacts and regalia. Symbols represent identifiable spiritual information associated with particular ancestral spirits that can be invoked to participate in healing and healing rituals. The sacred spiritual dressing in form of regalia contain spiritually significant materials from animal, plants and mineral kingdoms are imbued with spiritual powers. For example; long Kinganda robes *(kanzu),* head gear, animal hides, knotted bark-cloths, spears, armlets, wristlets, smoking pipe, walking stick among others, facilitate communication with ancestral spirits. One participant said *“the tree exudate/resin from Canarium schweinfurthii (Muwafu) tree, has special spiritual function, and when I put it on fire, its smoke assists me to communication with the dead people, ancestral spirits and the Creator. That is why it is used by various religions such as Muslims, Christians and us the traditionalists”* (Participant 7).

Participants expressed beliefs in symbolism of colors and associated ancestral spirits. The most symbolic colors are; white, black, red, shades of brown, golden-yellow and mixed colors. Although white is general for ancestral spirits, white and silver are specific for ancestral spirit Mukasa. Black is associated with spirit of dead person (*Muzimu*), while red is for Kiwanuka, golden-yellow for Musoke and shades of brown for Muwanga and their respective animals, birds, artefacts and regalia. Multi-colored animals and birds are associated with mystic ancestral spirits (*Misambwa*). Black is associated with bad luck, and white associated with good luck (*mukisa),* blessings (*bweeza)*, and fertility. Much of all these are also communicated through dreams.

### Beliefs in spiritual powers of dreams and associated messages

Participants believe in dreams and the contained spiritual messages. They claim that dreams are very informative if the communication is well understood as expressed by various healers. For example, *“During my health care practice, I get health care information through dreams, signs and actions”* (Participant <u>9).</u> *“ In some cases, I give medicine to my clients to facilitate them to dream which are then interpreted for meaning towards the required solution.”*(Participant <u>7).</u>

### Beliefs in spiritual powers generated through rituals

The traditional spiritual healers believe that a lot of healing spiritual powers are generated through thoughtful observance of individual and communal cultural rituals and understanding the spiritual significance of the associated elements such as fire and water, and plant and animal materials used during bathing, singing, offerings and sacrifice. Aerva lanata (*Lweza*), Momordica foetida (*Bombo),* and Dracaena fragrans *(Mulamula)* were most commonly used plants during spirits harmonization, empowerment and ritualistic training of ancestral spirits, and healing. Healers believe that rituals attract the functional presence of ancestral spirits, spiritual powers, and people into congregation (*lukiiko*). Rituals are effective for connection, communication, acknowledgement and to appease ancestral spirits and rejuvenate the spiritual powers of spirits. Furthermore, rituals are used to address physical, spiritual, moral, psychological, social and cultural issues during healing for individuals, family, relatives, friends, and community members. Rituals are used for cleansing, to remove bad spirits, treat witchcraft, and for prevention illness and social problems. One participant said that *“Katonda* (“God”) has authority over His creation. *I believe that when I pray to the Creator* (God) *and his creation including the spirits, I get the response I want.”* (Participant 9). The common communal rituals include sacrifices (*Kusaddaaka*), throat-cutting (*Kusalira*), communal offerings and meals (*Kugabula*).

### Sacrifice and throat-cutting, and communal meals and offerings

Sacrifice and throat-cutting of animals and birds of specified sex, maturity and colors, may be required during harmonization, rejuvenation and empowerment of ancestral spirits, and during healing. The sacrificed animals and birds may or may not be used in the communal meals and offerings. Communal meals (*ebijjulo*) and offerings (*kugabula*) to acknowledge and appease spirits are performed at specified sacred places, home, or guided by the ancestral spirits or knowledgeable ritual leader. The meals and offerings usually include fruits such as mangoes, pineapples, oranges, water melon, sugarcane, sweet bananas, pawpaw, guavas, and avocado among others. The drinks include alcohol, local brew, water, fresh banana juices, honey, and other juices as requested by the particular ancestral spirits. Others include cooked or roasted food and meat. The presence of Mukasa in any form, sign or symbol during or after the ritual affirms its success. One participant said, *“Mukasa appears physically or in signs to approve the perfection, rationality or significancy of most rituals performed successfully, even in simple rituals”* (Participant 8).

## Discussion

Study participants believed in ancestral spirits, spirit mediums, and spiritual powers contained in sacred places, living and non-livings materials, and non-materials such as dreams, symbols and rituals. Ancestral spirits are believed to have spiritual powers and are categorized into three; spirits of dead persons (*Mizimu*), mystical spirits (*Misambwa*) and assistant or worker spirits (*Mayembe*). Ancestral mystical spirits (*Misambwa*) were further grouped into three; water-related (*Mukasa, Kiwanuka* and *Musoke),* the royal (Ndawula, Kawumpuli and Bamweyana} and dryland (Ddungu and Muwanga) ancestral spirits. The head of the ancestral worker/assistant spirits was said to be Lubowa. Mentioned sacred places included natural forests, water bodies, shrines and fire places. The living materials with spiritual powers include plants, animals, birds, and reptiles. The non-living materials include artefacts, regalia and colors. The Sun, Moon, Stars, symbols and dreams were non-materials believed to be linked with spiritual powers. Furthermore, rituals, sacrifice, communal meals and offerings were connected with spiritual powers.

The central belief relates to the previous findings that health, illness and disease are controlled by non-human entities (41), and that out of cultural experience and personal expectations, people’s health require social relationships and engagements with nature and related spiritual powers (42). Various researchers have for long established that in Africa, the relationships between the living and their ancestors are believed to be mutually beneficial (43,44), and linked by spirit mediums (45,46). Furthermore, illness and associated suffering are socialized (21) through ritual healing and symbolic interpretations (20) in line with cultural, spiritual and supernatural causal relations (40,47–52).

According to Kleinman, every culture has its own particular explanations for health and illness, and its own culturally appropriate treatment approaches (53). For example, the worldview and perceptions of Konso people are linked to their local cosmology and traditions (20,54). People in many African ethnic groups recognize, respect, and relate to their ancestors as if the dead were still alive (44,51), but remain conscious that when angered, spiritual beings may cause illness and disease (22,55). Nonetheless, like elsewhere, the anger of ancestral spirits are usually appeased with prayers, rituals and offerings (56,57). Other studies reported that during healing, access to guidance by ancestral spirits is through dreams, diviners, and visions (44,58,59). According to Mokgobi, Africans give variable names to ancestral spirits. For example, the Bapedi, Batswana, and Basotho refer to them as ‘*badimo’*, the Amazulu call then *‘Amadlozi’* and the Amaxhosa call them *Iinyanya’* while the Baganda refer to them as *Mizimu*, *Balongo*, and *Misambwa* (60).

Beliefs in spiritual beings is not exclusive to African culture. Religious cultures such as Buddhism, Christianity, Hinduism, Islam, and Judaism all subscribe to beliefs in spiritual beings of some sort whom they implore for healing through actions, words, events and rituals (44,61). Although the context vary, Traditional Chinese Medicine pivots on the belief of a substance that is similar in all things and is symbolically connected to everything through “qi”, known as “vital force” in the West, which when out of balance results in illness (62). It is also noted that this “qi” substance is found in all natural formations including the Sun, Moon, Stars, the Earth, plants, animals, soils, and waters (62). Likewise, Ayurveda medicine in India, is based on traditional beliefs that the five elements (Pancha Bhuta) in the universe are also present in the human body, and are key to the connectivity of everything (63).

Rituals have been documented to reduce stress, strengthens resilience (64), help people to make sense of life-altering events (64,65), and heighten mutual interdependence among all creatures (19,66). Some India communities are reported to use words in rituals embedded with psycho-spiritual–socio-ecological cosmological worldview that emphasize family bonds, plants and animals as symbols of divinity, and link to spirits and ancestors while respecting cultural differences (67). In Pakistan, healing rituals engage many people and are associated with sacred activities to address physical and emotional issues (68).

On the other hand, the findings are in contrast to formal medical system which is based on scientific knowledge of biological and biochemical compounds that separates the body from the mind (69). Formal medical system is characterized by cell theory, germ theory, and bacteriology which seek to identify a causative pathogen, and that illness is always reducible to a physical, biological disease, or biomedical problem that can be medicalized (69).

In essence, African non-religious healing beliefs are linked with trance, ancestral communication and spiritual forces (70) that are celebrated by means of rituals involving offerings, animal sacrifice, oral narratives, cultural devotional songs (41), and various mandatory healing regimens (18). Currently, artefacts are used to reengage spiritual traditions (71), while other authors have advocated for re-contextualization of African cultural rituals and symbolism (72) for their collective African spiritual worldviews (7).

The main issue we are trying to highlight is that there is such vast informal practice within the communities that is seldom considered nor accounted for in the health service delivery or health systems discussions. These values as Kabir Sheikh puts it - “in addition to these concrete and tangible expressions of health systems, the “software”—by which we mean the ideas and interests, values and norms, and affinities and power that guide actions and underpin the relationships among system actors and elements—are also critical to overall health systems performance.”(73). These sociological findings are important aspects for considerations in public health and health policy formulations, given that traditional spiritual healers are part of the informal unregulated health care providers. Our contention in the article is that public health policy should be sensitive to this and the system has to have contextualized approaches in addressing these pathways or determinants to health.

This study is in line with earlier recommendations that the system of traditional healthcare providers should be studied and contextualized in ways of own (74,75). It also contributes to Articles 24 and 31 of the United Nations that encourage indigenous peoples to maintain, control, protect and develop their traditional medicine, health and practices and cultural expressions (76).

## Study limitations

The study participants were from 9 out of 56 clans of Baganda, 8 out 18 counties of Buganda and 11 out of 24 districts of Central Uganda, which limited the scope representation of Baganda clans and Buganda geographical in Central Uganda. It also limited the generation of data and more insightful conclusions.

## Conclusion

Ancestral spirits are central for the healing abilities of Baganda traditional spiritual healers (*Balubaale)*. *Balubaale* are characterized by beliefs and practices regarding the co-existence and linkage of human beings, nature, and spiritual beings. Ancestral spirits are believed to provide healing information to spiritual healers accessed through spirit possession, dreams, symbols and rituals.

This qualitative study provides key information on traditional beliefs underlying the use of ancestral spirits for healing among Baganda spiritual healers, and the spiritual world involving ancestral spirits and nature. The study adds to the limited existing knowledge, stimulates academic and non-academic discussions regarding ancestral spirits in health care, and provides a basis for broader examination of the link between ancestral spirits, traditional spiritual healers and public health in relation to policy considerations.

## Recommendations

We recommend further research studies involving greater numbers of representatives of more clans, counties and districts to concretize the findings. We also recommend to extend the study to other tribes, regions and districts in Uganda so as to compare the findings and contribute towards appropriate policy considerations.

## Data Availability

Data is available and is attached

## Acknowledgement

We extend our gratitude towards the leadership of Mbarara University and the PHARMBIOTRAC program for the financial support towards the PhD study of the first author. Particular thanks to Dr. Casim Tolo, Professor Patrick Ogwang and Engineer Anke Weisheit. We thank the study participants and their ancestral spirits for their committed participation in the study and for the information shared. We are equally grateful to the leaders of traditional healers’ associations, namely Sylivia Namutebi (Maama Fiina) of Uganda N’eddagala Ly’ayo N’obuwangwa Bwaffe, Musasizi Karim of NACOTHA, and Karim Walyabira of Uganda N’eddagala Ly’ayo. We acknowledge Oweky. Kyewalabye Male of Buganda Kingdom for introducing us to the clan leaders. We thank the Baganda clan leaders and district leadership for your support in accessing the study participants. We thank the research assistants Mr. Eric Kibirige Mukasa and Ms. Nambuya Barbra for the field work. We also thank Dr. John Chrysostom Katongole, Jovent K. Obbo of Bugema University and Dr. Kizito Simon of

